# Noradrenergic modulation of saccades in Parkinson’s disease

**DOI:** 10.1101/2024.01.04.24300879

**Authors:** Isabella F. Orlando, Frank H. Hezemans, Rong Ye, Alexander G. Murley, Negin Holland, Ralf Regenthal, Roger A. Barker, Caroline H. Williams-Gray, Luca Passamonti, Trevor W. Robbins, James B. Rowe, Claire O’Callaghan

## Abstract

Noradrenaline is a powerful modulator of cognitive processes, including action-decisions underlying saccadic control. Changes in saccadic eye movements are common across neurodegenerative diseases of ageing, including Parkinson’s disease. With growing interest in noradrenergic treatment potential for non-motor symptoms in Parkinson’s disease, the temporal precision of oculomotor function is advantageous to assess the effects of this modulation. Here we studied the effect of 40 mg atomoxetine, a noradrenaline reuptake inhibitor, in nineteen people with idiopathic Parkinson’s disease using a single dose, randomised double-blind crossover placebo-controlled design. Twenty-five healthy adult participants completed the assessments to provide normative data. Participants performed prosaccade and antisaccade tasks. The latency, velocity and accuracy of saccades, and resting pupil diameter, were measured. Increased pupil diameter on the drug confirmed its expected effect on the locus coeruleus ascending arousal system. Atomoxetine improved key aspects of saccade performance: prosaccade latencies were faster and the saccadic main sequence was normalised. These improvements were accompanied by increased antisaccade error rates on the drug. Together these findings suggest a shift in the speed-accuracy trade-off for visuo-motor decisions in response to noradrenergic treatment. Our results provide new evidence to substantiate a role for noradrenergic modulation of saccades, and based on known circuitry we advance the hypothesis that this reflects modulation at the level of the locus coeruleus–superior colliculus pathway. Given the potential for noradrenergic treatment of non-motor symptoms of Parkinson’s disease and related conditions, the oculomotor system can support the assessment of cognitive effects without limb-motor confounds on task performance.

---

The oculomotor system sits at the interface of a perception-action cycle that integrates visual information from the environment with an organism’s current internal state, facilitating the decisions underlying adaptive behaviour. Saccadic eye movements provide a means of exploring the visual world, subject to reflexive and high-order cognitive processes. Control of saccades involves the visual, prefrontal and parietal cortices, basal ganglia, thalamus, brainstem reticular formation and superior colliculus [1]. Distributed neural processes across this network enable interactions between an organism’s environment, goals and behaviour [2,3]. The oculomotor output of the superior colliculus integrates this exogenous and endogenous information, enabling optimal decisions about where and when to direct a saccade [4]. In this way, eye movements reflect the dynamics of decision processes, including information accumulation, evaluation, deliberation and choice [5].

Eye movement abnormalities are not a defining feature of Parkinson’s disease, but they are nonetheless commonly affected, with marked individual differences [6–10]. With progression of motor and cognitive impairment, prolonged saccadic latencies, fragmented, hypometric saccades and increased size and frequency of square wave jerks may be observed [10–13]. However, the latency and accuracy of saccades have been of particular interest in identifying the effect of disease and treatment on decision-making processes. For example, on more cognitively demanding tasks, including memory-guided saccades and antisaccades, inhibitory deficits are evident early in the illness and worsen with disease progression and cognitive impairment [13–17].

Saccade deficits in Parkinson’s disease are often interpreted in light of dopamine cell loss in the substantia nigra pars compacta. In Parkinson’s disease, reduced substantia nigra pars compacta dopaminergic input to the caudate results in a loss of inhibitory tone from the globus pallidus externus; this permits excessive outflow from the subthalamic nucleus to the substantia nigra pars reticulata, leading to tonic inhibition over the superior colliculus which controls the downstream premotor circuit for saccade initiation [9,18]. However, the diversity of saccade deficits and their heterogeneity among individuals with Parkinson’s disease suggests mechanisms extrinsic to the basal ganglia and beyond dopaminergic modulation. Noradrenergic contributions to the phenotype of Parkinson’s disease are of particular interest, because of therapeutic opportunities [19].

Noradrenergic projections from the locus coeruleus innervate intermediate layers of the superior colliculus [20–22]. In the non-human primate visual system, noradrenergic fibres target regions involved in spatial processing and visuomotor responses (i.e., the tecto-pulvinar-extrastriate visual structures) [22–24]. Other regions of the oculomotor network, including the thalamus, prefrontal, parietal and visual cortices receive dense noradrenergic innervation from the locus coeruleus [25,26]. Noradrenaline supresses spontaneous firing and enhances stimulus-evoked firing, producing a net increase in signal-to-noise ratio [27,28]. In sensory circuits, a “gating” effect is observed, where noradrenaline induces a robust cellular response to otherwise subthreshold visual stimuli [29,30].

The locus coeruleus is one of the earliest sites of pathology in Parkinson’s disease [31], with severe deficits in the symptomatic stages [32]. This noradrenergic deficit is implicated in diverse cognitive and neuropsychiatric symptoms [19,32–37]. In light of this, there is growing interest in noradrenergic treatment for non-motor symptoms in Parkinson’s disease and related disorders [38,39]. Oculomotor measures are particularly relevant in Parkinson’s disease, as they offer a means of assessing decision mechanisms and treatment effects without the confounding effect of limb akinetic-rigidity. Here we test the hypothesis that atomoxetine, a noradrenaline reuptake inhibitor, improves oculomotor control in Parkinson’s disease. We measured prosaccade and antisaccade task performance in people with Parkinson’s disease using a single dose double-blind, crossover, placebo-controlled design.

## Methods

This study was part of a broader project: study design and participant characteristics overlap with previous publications [36,37]. The study was approved by the local Ethics Committee (REC 10/H0308/34) and participants provided written informed consent.

### Participants

Nineteen people with Parkinson’s disease were recruited through the University of Cambridge Parkinson’s disease research clinic and Parkinson’s UK volunteer panels. All participants met United Kingdom Parkinson’s Disease Society Brain Bank diagnostic criteria, were aged between 50-80 years and had no contraindications to atomoxetine or 7T MRI. No participant met criteria for dementia [40]. Twenty-six age-, sex- and education-matched healthy control participants were recruited. Control participants were screened for a history of psychiatric or neurological disorders, and were not taking psychoactive medications. One control participant was excluded from all analyses due to inadequate eye tracking data.

### Study procedure

Participants with Parkinson’s disease were tested over three sessions. In the first session, they completed MRI scanning and clinical assessment of cognition and motor function (see Table 1). The second and third sessions comprised a double-blind, placebo-controlled crossover design, where participants were randomised to receive 40mg of atomoxetine or placebo. Session two and three visits were ≥ 6 days apart [mean = 7.4 days; standard deviation (SD) 1.8; range 6-14]. Blood samples were taken 2 hours post drug/placebo administration, corresponding to the predicted peak in plasma concentration [41]. Mean plasma concentration [42] was 264.07 ng/mL after atomoxetine (SD = 124.50 ng/mL, range: 90.92-595.11 ng/mL) and 0 ng/mL after placebo. Participants then commenced an experimental task battery including the oculomotor tasks reported here. Blood pressure and pulse rate were taken across the sessions, and visual analogue scales administered to monitor mood/arousal levels (see Supplementary Material). All sessions were completed at a similar time of day, with participants on their regular anti-parkinsonian medications. Control participants provided normative data, they were tested in a single session and did not undergo the drug manipulation.

**Table 1.** Data are presented as mean (standard deviation (SD)). Comparisons of patient and control groups were performed with independent samples t-tests or contingency tables as appropriate. MMSE = Mini-Mental State Examination [68]; MoCA = Montreal cognitive assessment [69]; ACE-R = revised Addenbrooke’s Cognitive Examination [70]; MDS-UPDRS = Movement Disorders Society Unified Parkinson’s Disease Rating Scale [71].

|  |  | **PD** | **Controls** | **BF** | ***p*** |
| --- | --- | --- | --- | --- | --- |
| Age (years) |  | 67.11 (7.05) | 65.40 (5.42) | 0.42 | .388 |
| Education (years) |  | 14.05 (2.27) | 14.64 (3.16) | 0.36 | .477 |
| Male / Female |  | 15 / 4 | 14 / 11 | 1.15 | .204 |
| MMSE |  | 29.47 (0.70) | 29.80 (0.50) | 1.10 | .093 |
| MoCA |  | 28.11 (1.76) | 28.52 (1.39) | 0.41 | .403 |
| ACE-R | Total Score | 94.89 (3.71) | 97.56 (3.23) | **3.65** | **.017** |
|  | Attention & Orientation | 17.84 (0.37) | 17.96 (0.20) | 0.62 | .225 |
|  | Memory | 23.68 (1.97) | 25.04 (1.18) | **7.65** | **.012** |
|  | Fluency | 12.00 (2.08) | 12.76 (1.61) | 0.63 | .196 |
|  | Language | 25.84 (0.50) | 25.88 (0.44) | 0.31 | .795 |
|  | Visuospatial | 15.63 (0.50) | 15.80 (0.65) | 0.43 | .333 |
| MDS-UPDRS | I: Nonmotor experiences | 9.00 (4.18) |  |  |  |
|  | II: Motor experiences | 12.63 (4.26) |  |  |  |
|  | III: Motor Examination | 28.42 (11.60) |  |  |  |
|  | IV: Motor Complications | 0.47 (0.96) |  |  |  |
|  | Total Score | 50.58 (17.20) |  |  |  |
| Hoehn and Yahr stage |  | 2.26 (0.45) |  |  |  |
| Disease duration (years) |  | 4.15 (1.72) |  |  |  |
| Levodopa equivalent daily dose (mg/day) |  | 644.55 (492.81) |  |  |  |

### Oculomotor assessments

Pupil size and eye movements were recorded using an EyeLink-1000 portable duo eye tracker at a sampling rate of 500 Hz (See Supplementary Material for full set up details). The tasks were programmed using Eyelink Experiment Builder software, with further preprocessing conducted in R (version 4.2.1, R Core Team, 2022). Our analyses used saccade detection, velocity and amplitude calculations from the standard Eyelink algorithms.

#### Resting pupil diameter

Pupil diameter was recorded over a 3-minute period at rest. Participants were asked to maintain fixation on a central crosshair. We first assessed signal quality by dividing each participant’s samples into 15 s time windows and removing windows with >15% of samples missing. Standard preprocessing followed: we removed 100 ms before and after blinks [44] and linearly interpolated the missing traces, then applied Hanning window smoothing [45]. Pupil diameter was calculated in arbitrary Eyelink units and z-scored. We removed extreme samples that were ±3 standard deviations from the mean and averaged across all surviving time windows.

#### Prosaccade and Antisaccade tasks

For the prosaccade and antisaccade tasks, participants were presented with a red circle (the target) on a black background. Each trial began with the target presented centrally for a duration varying between 800-1200 ms. The target stimulus then disappeared and following a 200 ms gap it appeared on the left- or right-hand side of the screen. The target had six possible positions on the horizontal axis, presented in a randomised order. From the centre (0°), target locations were ±3°, ±5°, ±7°. Both tasks included 24 trials. In the prosaccade task, participants were instructed to direct their gaze towards the target. In the antisaccade task, participants were instructed to direct their gaze away from the target to the other (blank) side of the screen.

For analysis, in each trial we identified the primary saccade – i.e., the first valid saccade following the target’s appearance on either side of the screen [46,47]. The primary saccade was defined by an amplitude between >1.5° and < 10° (i.e., to rule out microsaccades and saccades too extreme to remain on the screen [48,49]) and a latency between > 90 ms and < 2500 ms (i.e., to rule out anticipatory saccades [50–52] and inattention [53]).

Prosaccade analysis was conducted on the first valid saccade directed towards the target. Antisaccade analysis was performed on the first valid saccade regardless of direction, and identified as a correct antisaccade if the direction was opposite to the target. Prosaccade trials with latencies, peak velocities or amplitudes more extreme than ±3 standard deviations from the participant’s mean were removed; similarly, this was done for antisaccade trials with respect to latency.

For prosaccades we measured velocity and amplitude throughout the saccade, by normalising the duration of each saccade and then dividing into 10 time bins. For each prosaccade we measured response latency (i.e., time from target display to initiation of the primary saccade), peak velocity (i.e., maximum velocity during the saccade) and absolute amplitude (i.e., angular distance travelled during the saccade). There is a well-established relationship known as the “main sequence” where a saccade’s velocity increases as amplitude increases [54]. To account for this, we used the peak velocity residuals as our velocity measure of interest, calculated by regressing amplitude scores against velocity scores [55–58]. For the antisaccade task we calculated error rates as the proportion of incorrectly directed antisaccades from the total completed trials; we examined latency for both correct and incorrect antisaccades.

To further explore the main sequence, we fitted a square root model [59], which is a robust model to quantify the main sequence in smaller sample sizes [60]:

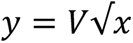

Where *y* is the peak velocity, *x* is the absolute amplitude and *V* is the coefficient [60–62]. 1000 bootstrapped estimates of *V* were simulated for each saccade, from which the median *V* was computed [62]. To determine how the placebo and atomoxetine groups deviated from the controls’ main sequence, for each observed peak velocity value in the patients we subtracted the modelled velocity value at a matched amplitude, using the curve derived from controls [62] (illustrated in Figure 2b). This created a Δ peak velocity value, where higher or lower values reflect greater deviation from the normative model.

### Statistical analysis

For all oculomotor measures we compared people with Parkinson’s disease on atomoxetine *versus* placebo. Separately, we compared participants with Parkinson’s disease on placebo *versus* controls. Analyses were conducted in R (version 4.2.1, R Core Team, 2022). We applied linear mixed models constructed using trial-wise data, accounting for repeated measures and pseudoreplication using random effects with a fixed effect of visit order also included. Frequentist models were implemented using the ‘afex’ package [63] and the ‘emmeans’ package [64] for *post hoc* comparisons with Sidak adjustments for multiple testing. Bayes factor (BF) analyses were implemented using ‘BayesFactor’ [65] and ‘bayestestR’ [66] packages. We report the Bayes factor (BF) for the alternative hypothesis over the null hypothesis (i.e., BF_10_), with BF > 1 indicating relative evidence for the alternative hypothesis and BF > 3 indicating positive evidence for the alternative hypothesis; and BF < 0.33 indicating positive evidence for the null hypothesis [67].

### Data availability

Code and data to reproduce figures and statistical analyses are available through the Open Science Framework (OSF_link_to_be_added).

## Results

### Demographics and clinical characteristics

Parkinson’s disease and control groups were well matched on demographics, but the patient group scored significantly lower on the ACE-R and its memory subscale (Table 1).

### Resting pupil diameter

Mean pupil diameter was significantly increased on atomoxetine compared to placebo [(*F*(1, 15) = 16.68, *p* < 0.001, BF = 34.46)] (Figure 1a). Mean pupil diameter in the patient-placebo group was not significantly different from controls [(*F*(1, 40) = 0.43, *p* = 0.517, BF = 0.364)].

**Figure 1.**
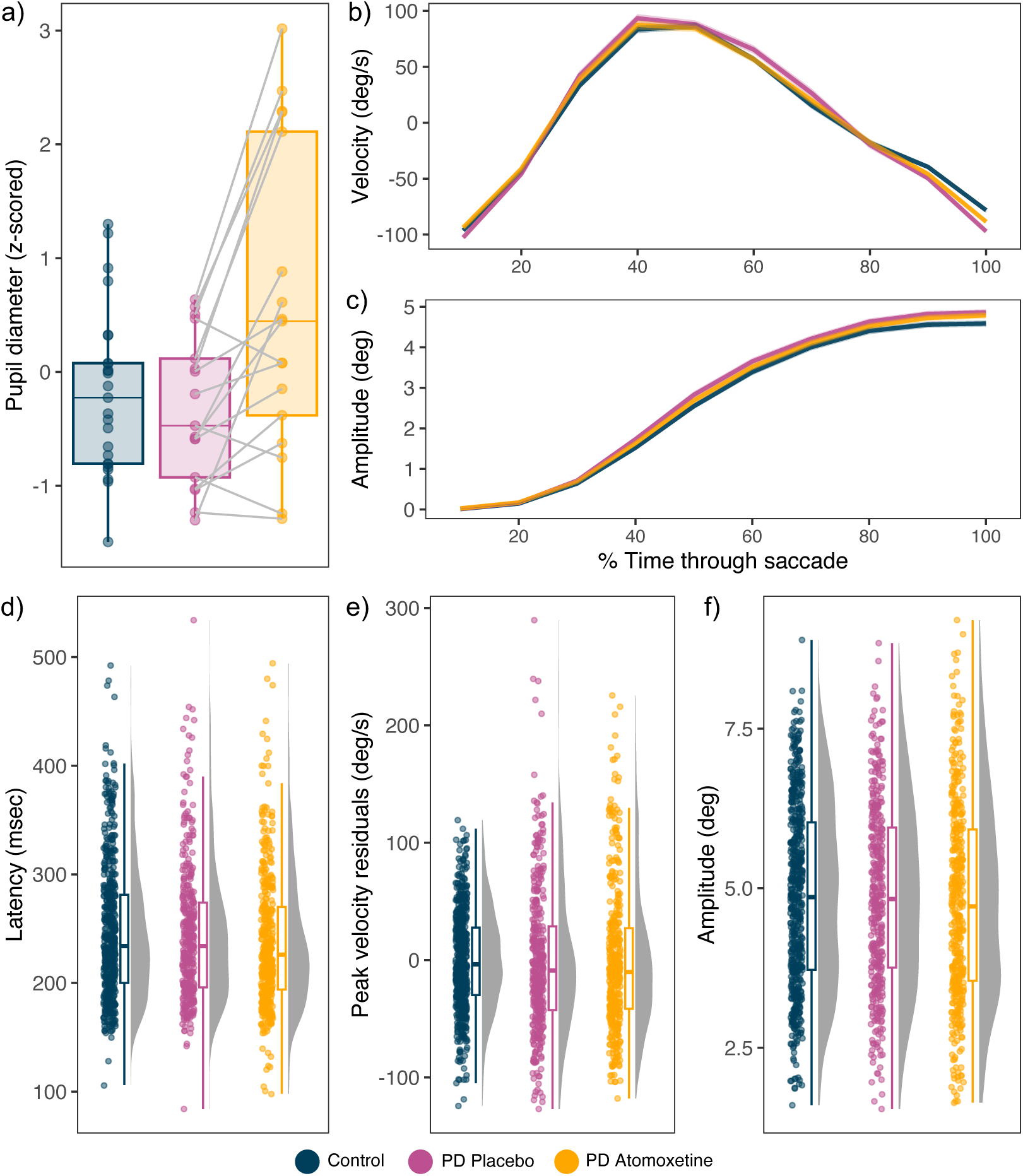
a) Mean z-scored pupil diameter at rest, with grey lines representing within subject comparisons. b) Mean velocity residuals and c) Amplitude throughout the time course of prosaccades, where the x-axis is % of normalised time with 0 indicating the start of a saccade and 100 indicating the end. Shaded ribbons represent standard error of the mean. d) Latency, e) Peak velocity residuals, and f) Amplitude of prosaccades for the three groups. Data points show trial-wise data, representing the primary saccade for every trial per participant.

**Figure 2.**
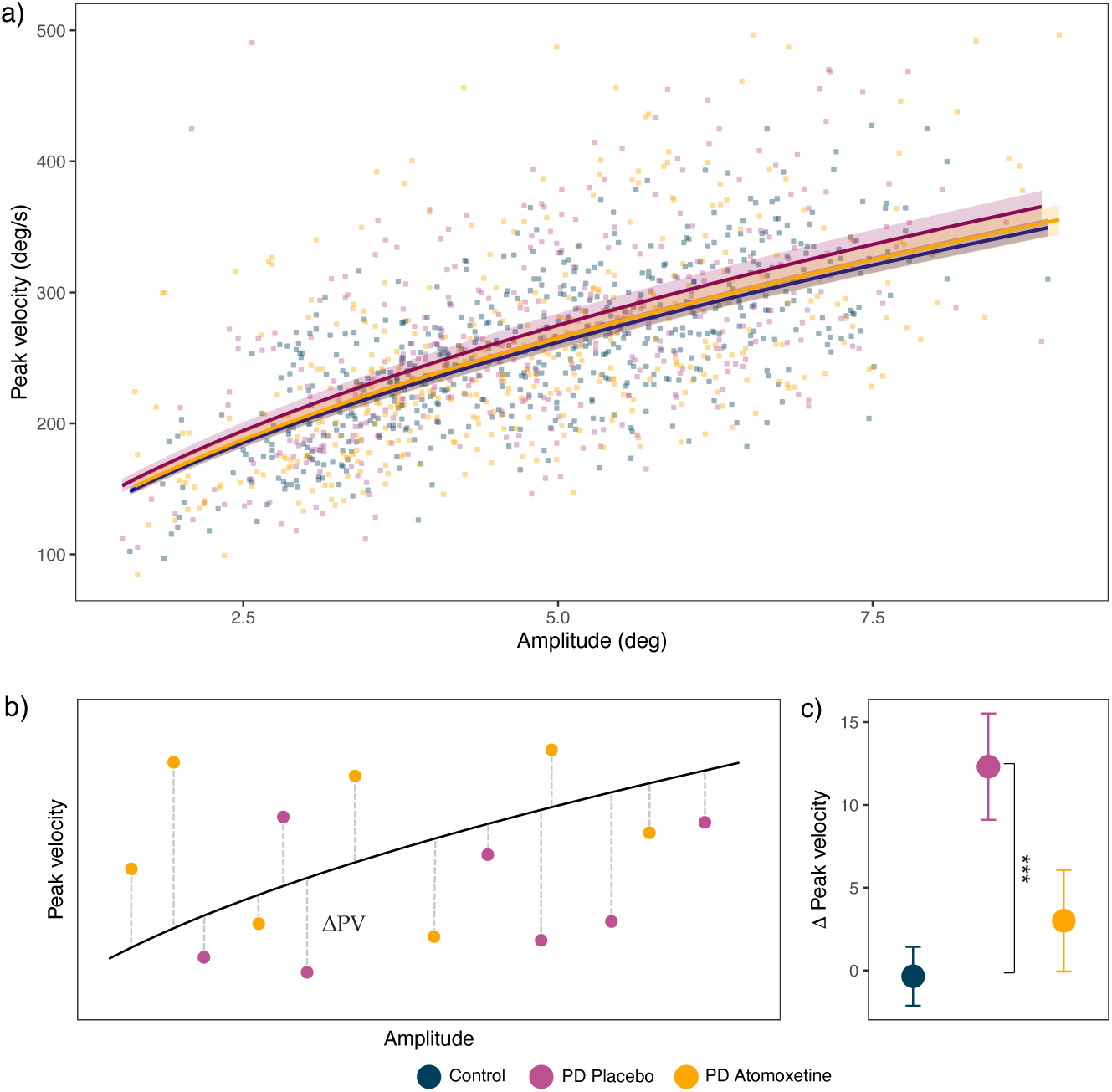
a) A square root model was fit to estimate the main sequence of prosaccades in each group. Solid lines illustrate the median fit computed over 1000 bootstrapped estimates of the coefficient (*V*), with 95% confidence intervals displayed by the shaded ribbons. b) Schematic of the peak velocity delta (Δ) calculations for each primary prosaccade from the normative main sequence curve (control curve estimated in Figure a). c) Mean Δ peak velocity for the three groups with standard error bars.

### Prosaccade velocity and amplitude over the saccade time course

All groups demonstrated the expected velocity and amplitude trajectories throughout prosaccades, with a symmetric profile of acceleration and deceleration around the peak velocity (Figures 1b, 1c). In the Parkinson’s disease group, prosaccade mean velocity residuals and mean amplitude across 10 time bins were not significantly different after atomoxetine versus placebo. There was no interaction between drug condition and time bin, and no effect of visit. Comparing the Parkinson’s disease group on placebo with control participants, there was no significant difference in mean velocity and amplitude across the time course, with no interaction between group and time bin (Results detailed in Supplementary Material).

### Prosaccade latency

There was evidence of faster saccadic responses on atomoxetine (Figure 1d), with significantly reduced latencies compared to placebo [*F*(1, 13.91) = 4.64, *p* = 0.049, BF = 2.13] We note that the BF was below conventional threshold for positive evidence (i.e., BF > 3), so this effect should be regarded as “anecdotal”. The effect of visit order was not significant (*F*(1, 13.55) = 3.89, *p* = 0.069, BF = 1.16). Latency did not differ between the Parkinson’s disease group on placebo and control participants [*F*(1, 41.88) = 0.050, *p* = 0.828, BF = 0.213].

### Prosaccade peak velocity and absolute amplitude

For peak velocity residuals (Figure 1e), there was evidence for no difference on atomoxetine compared to placebo [*F*(1, 16.59) = 0.01, *p* = 0.922, BF = 0.071]. For absolute saccade amplitude (Figure 1f), there was also evidence for no difference between the conditions [*F*(1, 16.47) = 0.47, *p* = 0.502, BF = 0.321]. With target location included as a covariate to account for the three target distances, there was a main effect of location [*F*(2, 30.99) = 173.59, *p* < 0.001, BF = 2.42 × 10^119^] reflecting larger amplitudes with increasing target distance; there was no interaction between target location and drug condition (*F*(2, 35.06) = 0.19, *p* = 0.826, BF = 0.037). For both peak velocity and absolute amplitude, there was no significant effect of visit order [peak velocity: (*F*(1, 16.59) = 0.19, *p* = 0.673, BF = 0.365); amplitude: *F*(1, 16.11) = 0.18, *p* = 0.674, BF = 0.092)].

The Parkinson’s disease group on placebo and controls did not differ in peak velocity residuals [*F*(1, 42.12) = 0.01, *p* = 0.958, BF = 0.403] or absolute amplitude [*F*(1, 41.25) = 0.04, *p* = 0.852, BF = 0.265]. There was the expected effect of target location on amplitude [*F*(2, 76.68) = 534.13, *p* < 0.001, BF = 1.67 × 10^220^] with no interaction between group and target location [*F*(2, 76.68) = 2.13, *p* = 0.126, BF = 0.847].

### Saccade main sequence

To determine the effect of atomoxetine on the main sequence, trial-wise model coefficients (*V*) for each prosaccade were analysed from square root models fitted to each group (Figure 2a). A significant main effect of the drug was found, suggesting atomoxetine alters the main sequence in Parkinson’s disease [*F*(1, 15.89) = 7.46, *p* = 0.015, BF = 3.33], with no effect of visit order [*F*(1, 15.48) = 0.06, *p* = 0.809, BF = 0.167]. The main sequence coefficient differed significantly between the Parkinson’s placebo group and controls [*F*(1, 40.40) = 12.64, *p* < 0.001, BF = 3.65]. One-sided t-tests tested whether Δ peak velocity values (i.e., the difference between the Parkinson’s disease groups’ observed values and the controls’ modelled main sequence curve) in the atomoxetine and placebo groups differed from zero (Figure 2b). Δ peak velocity values in the placebo group were significantly different from zero [*t* (379) = 3.83, *p* < 0.001, BF = 73.39], but there was evidence for no difference for the atomoxetine group [*t* (407) = 0.978, *p* = 0.328, BF = 0.09] (Figure 2c). Together, suggesting that atomoxetine normalised the main sequence to more closely resemble that of controls.

### Antisaccade errors

The proportion of errors in the antisaccade task (Figure 3b) was increased under atomoxetine compared to placebo [*F*(1, 16) = 7.57, *p* = 0.014, BF = 3.71], with no effect of visit order [*F*(1, 16) = 0.14, *p* = 0.715, BF = 0.419]. Errors did not differ between the Parkinson’s disease group on placebo and control participants [*F* (1, 41) = 0.738, *p* = 0.395, BF = 0.407].

**Figure 3.**
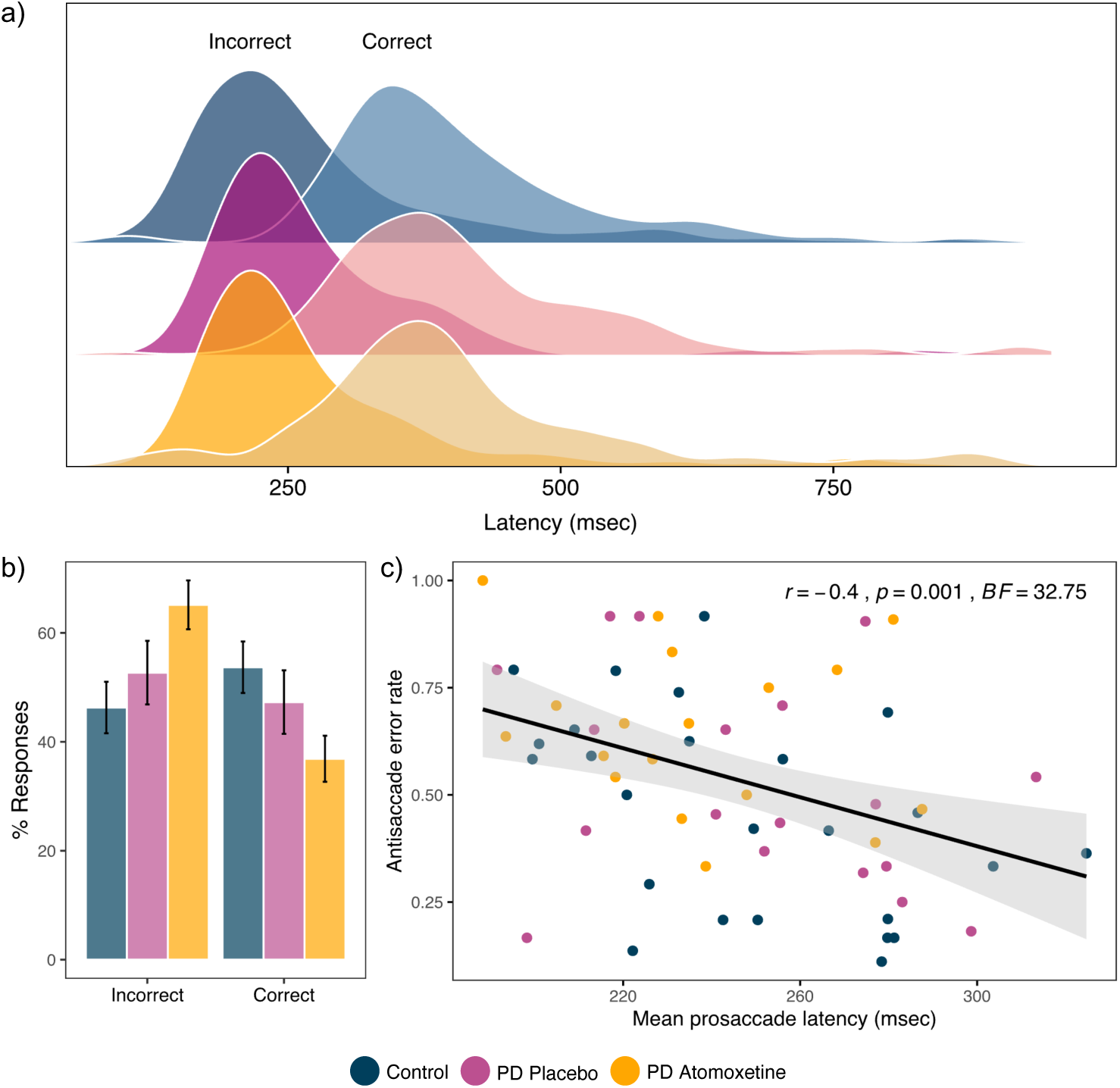
a) Antisaccade task latencies for incorrect (darker distribution on the left) and correct (transparent distribution on the right) trial types. b) Antisaccade task % of responses for incorrect and correct trial types. c) Correlation between mean prosaccade latency (msec) and antisaccade error rate for each participant coloured by condition.

Consistent with antisaccade errors reflecting impulsive responses, when latency was examined with correct vs. incorrect trial type included as a covariate, response latencies were significantly faster for incorrect antisaccade trials compared to correct trials (Figure 3a). This was the case for both atomoxetine vs. placebo [*F*(1, 15.87) = 58.12, *p* < 0.001, BF = 1.25 × 10^4^] and placebo vs. control [*F*(1, 37.55) = 105.60, *p* < 0.001, BF = 1.12 × 10^10^], with no interaction between trial type and condition [atomoxetine vs. placebo: (*F*(1, 20.47) = 0.62, *p* = 0. 438, BF = 0.161); placebo vs. control: (*F*(1, 37.55) = 0.33, *p* = 0.569, BF = 0.248)].

### Relationship between prosaccade latency and antisaccade error rate

That atomoxetine improved a measure on prosaccades (i.e., latency) while making antisaccade errors worse is consistent with the established relationship such that prosaccade latency is a reliable predictor of antisaccade error rate [72,73]. We confirmed this relationship via a Pearson’s correlation across the three groups pooled, which showed a significant negative correlation between mean prosaccade latency and antisaccade error rate (r = -0.402, *p* = 0.001, BF = 32.75; Figure 3c), demonstrating that individuals with a faster prosaccade latency made more errors on the antisaccade task.

#### Saccade performance on placebo in relation to disease severity and cognitive impairment

Given the literature showing that saccade deficits in Parkinson’s disease worsen with disease progression and cognitive impairment, we tested whether prosaccade latency, antisaccade error rate or main sequence deviation (i.e., Δ peak velocity values from controls’ main sequence) in the placebo group correlated with global cognition (ACE-R total score) or disease severity (MDS-UPDRS-III score). We conducted Pearson correlations with FDR correction applied within both families of comparisons. Both prosaccade latency and main sequence deviation showed a positive correlation with disease severity, consistent with worsening impairment with disease progression (latency: r = 0.549, *p*_(adjust)_ = 0.022, BF = 4.81; Δ peak velocity: r = 0.692, *p*_(adjust)_ = 0.003, BF = 29.89). (See Supplementary Material for full results and scatter plots).

## Discussion

The noradrenaline reuptake inhibitor atomoxetine improved several aspects of saccade behaviour, accompanied by an increase in antisaccade errors. These findings highlight a role for noradrenergic modulation of oculomotor control in Parkinson’s disease, and emphasise a key role for noradrenaline in modulating behavioural decisions, including eye-movements. In the context of potential noradrenergic treatments, our findings suggest that oculomotor control may be a useful surrogate of cognitive efficacy.

Atomoxetine had significant effects on the initiation of saccades: faster prosaccade latencies and more antisaccade errors were seen under atomoxetine relative to placebo. Consistent with speed and accuracy trade-off on a shared dimension, we found that higher rates of antisaccade errors were correlated with faster prosaccade latencies. Atomoxetine influenced this trade-off in individuals’ exertion of cognitive control, unconfounded by limb-motor deficits in the kinetics of execution of decisions.

At the heart of saccade control is a negotiation between salient stimuli that trigger automatic responses (e.g., a saccade to the stimulus), versus deliberative control (e.g., an antisaccade). Even a seemingly simple prosaccade towards a target occurs at approximately 200 millisecond latency in humans – a delay that is much slower than the constraints imposed by anatomy [74,75]. This has been called the “oculomotor procrastination” [76], in deciding not only where to look, but whether it’s worth looking. For antisaccade tasks, this can be conceptualised as a competition between two processes: supressing a prepotent response towards the target and generating a voluntary saccade in the opposite direction [1,77]. The balance between issuing a prepotent response versus exerting deliberative control is modulated by noradrenaline. This is most clearly demonstrated by cross-species evidence from the stop-signal task, where atomoxetine improves the ability to inhibit a prepotent motor response [78,79]. In a related domain, noradrenaline is implicated in the ability to shift responding from an established attentional set to a new set of responses [80–82]. In this way, noradrenergic modulation across different timescales influences the ability to flexibly reconfigure behavioural responses based on changing environmental contingencies. In our Parkinson’s disease patients on atomoxetine an invigorated response to the visual target, as evidenced by increased prosaccade latency, may come at the expense of implementing the cognitive control required to support antisaccades.

Noradrenergic modulation by atomoxetine may act via the locus coeruleus projections to the superior colliculus; or to cortical control systems (given the sparsity of noradrenergic terminals in the striatum). At the level of the locus coeruleus, systemic atomoxetine is associated with an increased phasic-to-tonic firing rate in response to sensory stimuli [83]. This will affect activity in coereuleal noradrenergic projections to the intermediate layers of the superior colliculus, which project in turn to the paramedian pontine reticular containing burst neurons responsible for initiating and stopping saccades [1]. The faster latencies and higher velocities of saccadic responses typically seen with increased arousal [84] may be mediated in part by elevated noradrenergic activity in the locus coeruleus pathway projecting to the intermediate superior colliculus [85]. In our patient group we found increased pupillary diameter on atomoxetine compared to placebo, corroborating the drug’s effect on an index of the ascending arousal system that has been closely linked to locus coeruleus activity [86].

Atomoxetine may also modulate cognitive control mediated by cortical networks. Descending inputs from the dorsolateral prefrontal cortex, frontal eye fields, supplementary eye fields and parietal eye fields contribute to “top-down” control over saccade performance [1]. Activity in these regions has been linked with successful inhibition on antisaccade trials [87] via both direct and indirect projections to the superior colliculus [88,89]. The dorsolateral prefrontal cortex is especially sensitive to the effects of noradrenergic modulation, with a non-linear “inverted-U” dose-response effect [90,91]. This non-linear response is offset by Parkinson’s disease, but to a different degree across different brain regions [92]. This means that a given oral dose of atomoxetine may optimise decisions mediated by one region while simultaneously driving decision-making processes at another region out of the optimal range. Therefore, even though the same dose improved limb-motor response inhibition in some individuals with Parkinson’s disease [34,36,93], it impairs antisaccade accuracy in this study, highlighting that optimal levels of noradrenergic modulation are often task-specific.

Another effect of atomoxetine was on the main sequence, the well-established relationship between a saccade’s amplitude and its velocity. In patients on atomoxetine the main sequence trajectory was more closely aligned with that of controls and was the only metric that differed significantly between participants with Parkinson’s disease on placebo versus controls. In our mild-moderate patient cohort a lack of group differences from controls is consistent with the heterogeneous findings in these saccade parameters in early-stage Parkinson’s disease [12,94]. In line with this, some features showed a relationship with disease severity: the extent of main sequence deviation from controls and slower latencies both increased with disease severity.

The main sequence describes a highly stereotyped relationship between the amplitude and velocity of saccades, with a linear increase for short saccades that asymptotes as speed saturates for larger saccades [54,60]. This non-linear property optimises the speed-accuracy trade-off, reducing the impact of motor noise on faster movements [95] and thus minimising the variability in saccade endpoints [96]. The superior colliculus is an upstream source of these non-linear kinematics [97–99]. Saccade related neurons in the intermediate superior colliculus are organised with rostral-caudal topography that corresponds to the amplitude and spatial location of the saccades they generate [100]. This arrangement is mirrored by a gradient of burst firing properties along the rostral-caudal extent, where distinct firing profiles occur for smaller to larger amplitudes – an organisation that could support non-linear dynamics [97]. Despite its stereotypy, the main sequence has been shown to deviate as a function of arousal [84,101], motivation [58] and in neurological disease [102–106]. Our results raise the novel possibility that the main sequence is sensitive to noradrenergic modulation, which we speculate is mediated by a direct locus coeruleus to superior colliculus pathway.

Atomoxetine may also increase dopamine in the neocortex, due to the paucity of dopamine transporters extracellular dopamine uptake is largely mediated by the noradrenaline transporter [107,108]. However, mediation of the effects of atomoxetine by cortical dopamine is unlikely: meta-analyses confirm prolonged latency on (versus off) dopamine medication, with no significant effects on velocity or amplitude parameters [109], and antisaccade error rate is not affected by dopamine medication [110]. Furthermore, atomoxetine is unlikely to strongly modulate the primary dopaminergic circuitry implicated in Parkinson’s disease oculomotor deficits, i.e., the basal ganglia pathways that drive substantia nigra inhibition over the superior colliculus, as the noradrenaline transporter is not responsible for significant dopamine reuptake in the striatum [107]. Together, this past literature argues against a dopaminergic interpretation of our results.

In summary, we demonstrate an effect of atomoxetine on saccadic control, with faster prosaccade latencies and a normalised main sequence trajectory, concurrent with more antisaccade errors which we suggest may reflect disrupted cognitive control from the frontal cortex. These findings highlight a noradrenergic role for oculomotor function relevant to oculomotor decision-making in Parkinson’s disease. Our findings highlight that basic oculomotor function may be a target for noradrenergic modulation in these conditions or provide an additional means of monitoring the effects of noradrenergic interventions. Noradrenergic drugs have potential to address cognitive and behavioural symptoms in Parkinson’s disease and related disorders.

## Data Availability

Code and data to reproduce figures and statistical analyses are available through the Open Science Framework.

## Acknowledgements

We thank all volunteers and their families for their participation, all staff at the Wolfson Brain Imaging Centre and NIHR Cambridge Clinical Research Facility for their help with data collection. We thank Sam Hutton for valuable advice on the task set up and analysis.

## Author contributions

C.O., F.H.H., J.B.R. and T.W.R. conceptualised and designed the project. C.O., F.H.H., R.Y., A.G.M., N.H., R.R. and L.P. contributed to the acquisition of data. I.F.O and C.O performed analysis and wrote the manuscript. All authors were involved in the interpretation of the data and with the revision of the final manuscript.

## Funding

This study was supported by Parkinson’s UK (K-1702) and the Cambridge Centre for Parkinson-Plus. F.H.H. was supported by a Cambridge Trust Vice-Chancellor’s Award and Fitzwilliam College Scholarship. N.H. was supported by the Association of British Neurologists-Patrick Berthoud Charitable Trust (RG99368). A.G.M. was supported by the Holt Fellowship (RG86564). C.H.W-G. was supported by a RCUK/UKRI Research Innovation Fellowship awarded by the Medical Research Council (MR/R007446/1). J.B.R. is funded from the Welcome Trust (103838; 220258), the Medical Research Council (MC_UU_00030/14; MR/T033371/1), the National Institute for Health Research Cambridge Biomedical Research Centre (NIHR203312: BRC-1215-20014); and a James S. McDonnell Foundation 21st Century Science Initiative Scholar Award in Understanding Human Cognition. C.O. was supported by a University of Sydney Robinson Fellowship and an Australian National Health and Medical Research Council EL2 Fellowship (2016866). This study was carried out at/supported by the NIHR Cambridge Clinical Research Facility and supported by the NIHR Cambridge Biomedical Research Centre (NIHR203312). The views expressed are those of the authors and not necessarily those of the NHS, the NIHR or the Department of Health and Social Care. For the purpose of open access, the authors have applied a CC BY public copyright licence to any Author Accepted Manuscript version arising from this submission.

## Conflict of Interest

The authors repost no conflicts of interest.

## Notes

### Competing Interest Statement

The authors have declared no competing interest.

### Clinical Trial

ISRCTN46299660

### Author Declarations

The study was approved by the Health Research Authority East of England-Cambridge Central Research Ethics Committee (REC 10/H0308/34). The trial was registered on ISRCTN registry with study ID ISRCTN46299660 (https://doi.org/10.1186/ISRCTN46299660). The study was retrospectively registered because it was exempt from Clinical Trials status by the UK Medicines and Healthcare Products Regulatory Authority (MHRA). However, we have endeavoured to retrospectively register it in order to meet compliance requirements for preprint and journal submissions.

## References

1. Munoz DP, Everling S. Look away: the anti-saccade task and the voluntary control of eye movement. Nat Rev Neurosci. 2004;5:218–228.

2. Fecteau JH, Munoz DP. Salience, relevance, and firing: a priority map for target selection. Trends in Cognitive Sciences. 2006;10:382–390.

3. Trappenberg TP, Dorris MC, Munoz DP, Klein RM. A Model of Saccade Initiation Based on the Competitive Integration of Exogenous and Endogenous Signals in the Superior Colliculus. Journal of Cognitive Neuroscience. 2001;13:256–271.

4. Merker B. The efference cascade, consciousness, and its self: naturalizing the first person pivot of action control. Frontiers in Psychology. 2013;4.

5. Spering M. Eye Movements as a Window into Decision-Making. Annual Review of Vision Science. 2022;8:427–448.

6. Anderson TJ, MacAskill MR. Eye movements in patients with neurodegenerative disorders. Nat Rev Neurol. 2013;9:74–85.

7. Antoniades CA, Spering M. Eye movements in Parkinson’s disease: from neurophysiological mechanisms to diagnostic tools. Trends in Neurosciences. 2023;0.

8. Kassavetis P, Kaski D, Anderson T, Hallett M. Eye Movement Disorders in Movement Disorders. Movement Disorders Clinical Practice. 2022;9:284–295.

9. Pretegiani E, Optican LM. Eye Movements in Parkinson’s Disease and Inherited Parkinsonian Syndromes. Frontiers in Neurology. 2017;8:592.

10. Brien DC, Riek HC, Yep R, Huang J, Coe B, Areshenkoff C, et al. Classification and staging of Parkinson’s disease using video-based eye tracking. Parkinsonism & Related Disorders. 2023:105316.

11. MacAskill MR, Graham CF, Pitcher TL, Myall DJ, Livingston L, van Stockum S, et al. The influence of motor and cognitive impairment upon visually-guided saccades in Parkinson’s disease. Neuropsychologia. 2012;50:3338–3347.

12. Mosimann UP, Müri RM, Burn DJ, Felblinger J, O’Brien JT, McKeith IG. Saccadic eye movement changes in Parkinson’s disease dementia and dementia with Lewy bodies. Brain. 2005;128:1267–1276.

13. Terao Y, Fukuda H, Yugeta A, Hikosaka O, Nomura Y, Segawa M, et al. Initiation and inhibitory control of saccades with the progression of Parkinson’s disease – Changes in three major drives converging on the superior colliculus. Neuropsychologia. 2011;49:1794–1806.

14. Antoniades CA, Demeyere N, Kennard C, Humphreys GW, Hu MT. Antisaccades and executive dysfunction in early drug-naive Parkinson’s disease: The discovery study. Movement Disorders. 2015;30:843–847.

15. Chan F, Armstrong IT, Pari G, Riopelle RJ, Munoz DP. Deficits in saccadic eye-movement control in Parkinson’s disease. Neuropsychologia. 2005;43:784–796.

16. van Stockum S, MacAskill M, Anderson T, Dalrymple-Alford J. Don’t look now or look away: Two sources of saccadic disinhibition in Parkinson’s disease? Neuropsychologia. 2008;46:3108–3115.

17. Zhang J, Rittman T, Nombela C, Fois A, Coyle-Gilchrist I, Barker RA, et al. Different decision deficits impair response inhibition in progressive supranuclear palsy and Parkinson’s disease. Brain. 2016;139:161–173.

18. Terao Y, Fukuda H, Ugawa Y, Hikosaka O. New perspectives on the pathophysiology of Parkinson’s disease as assessed by saccade performance: A clinical review. Clinical Neurophysiology. 2013;124:1491–1506.

19. Holland N, Robbins TW, Rowe JB. The role of noradrenaline in cognition and cognitive disorders. Brain. 2021;144:2243–2256.

20. Edwards SB, Ginsburgh CL, Henkel CK, Stein BE. Sources of subcortical projections to the superior colliculus in the cat. Journal of Comparative Neurology. 1979;184:309– 329.

21. Li L, Feng X, Zhou Z, Zhang H, Shi Q, Lei Z, et al. Stress Accelerates Defensive Responses to Looming in Mice and Involves a Locus Coeruleus-Superior Colliculus Projection. Current Biology. 2018;28:859–871.e5.

22. Morrison JH, Foote SL. Noradrenergic and serotoninergic innervation of cortical, thalamic, and tectal visual structures in old and new world monkeys. Journal of Comparative Neurology. 1986;243:117–138.

23. Foote SL, Berridge CW. New developments and future directions in understanding locus coeruleus – Norepinephrine (LC-NE) function. Brain Research. 2019;1709:81– 84.

24. Foote SL, Morrison JH. Extrathalamic Modulation of Cortical Function. Annual Review of Neuroscience. 1987;10:67–95.

25. Samuels ER, Szabadi E. Functional neuroanatomy of the noradrenergic locus coeruleus: Its roles in the regulation of arousal and autonomic function Part I: Principles of functional organisation. Current Neuropharmacology. 2008;6:235–253.

26. Schwarz LA, Luo L. Organization of the locus coeruleus-norepinephrine system. Curr Biol. 2015;25:R1051–R1056.

27. Berridge CW, Waterhouse BD. The locus coeruleus–noradrenergic system: modulation of behavioral state and state-dependent cognitive processes. Brain Research Reviews. 2003;42:33–84.

28. Foote SL, Freedman R, Oliver AP. Effects of putative neurotransmitters on neuronal activity in monkey auditory cortex. Brain Research. 1975;86:229–242.

29. Navarra RL, Clark BD, Zitnik GA, Waterhouse BD. Methylphenidate and atomoxetine enhance sensory-evoked neuronal activity in the visual thalamus of male rats. Experimental and Clinical Psychopharmacology. 2013;21:363–374.

30. Waterhouse BD, Sessler FM, Jung-Tung C, Woodward DJ, Azizi SA, Moises HC. New evidence for a gating action of norepinephrine in central neuronal circuits of mammalian brain. Brain Research Bulletin. 1988;21:425–432.

31. Braak H, Tredici KD, Rüb U, de Vos RAI, Jansen Steur ENH, Braak E. Staging of brain pathology related to sporadic Parkinson’s disease. Neurobiology of Aging. 2003;24:197–211.

32. Ye R, O’Callaghan C, Rua C, Hezemans FH, Holland N, Malpetti M, et al. Locus Coeruleus Integrity from 7 T MRI Relates to Apathy and Cognition in Parkinsonian Disorders. Movement Disorders. 2022;37:1663–1672.

33. Orlando IF, Shine JM, Robbins TW, Rowe JB, O’Callaghan C. Noradrenergic and cholinergic systems take centre stage in neuropsychiatric diseases of ageing. Neuroscience & Biobehavioral Reviews. 2023;149:105167.

34. Ye Z, Altena E, Nombela C, Housden CR, Maxwell H, Rittman T, et al. Improving Response Inhibition in Parkinson’s Disease with Atomoxetine. Biological Psychiatry. 2015;77:740–748.

35. Kehagia AA, Housden CR, Regenthal R, Barker RA, Müller U, Rowe J, et al. Targeting impulsivity in Parkinson’s disease using atomoxetine. Brain. 2014;137:1986–1997.

36. O’Callaghan C, Hezemans FH, Ye R, Rua C, Jones PS, Murley AG, et al. Locus coeruleus integrity and the effect of atomoxetine on response inhibition in Parkinson’s disease. Brain. 2021;144:2513–2526.

37. Hezemans FH, Wolpe N, O’Callaghan C, Ye R, Rua C, Jones PS, et al. Noradrenergic deficits contribute to apathy in Parkinson’s disease through the precision of expected outcomes. PLOS Computational Biology. 2022;18:e1010079.

38. David MCB, Giovane MD, Liu KY, Gostick B, Rowe JB, Oboh I, et al. Cognitive and neuropsychiatric effects of noradrenergic treatment in Alzheimer’s disease: systematic review and meta-analysis. J Neurol Neurosurg Psychiatry. 2022. 1 June 2022. 10.1136/jnnp-2022-329136.

39. Levey AI, Qiu D, Zhao L, Hu WT, Duong DM, Higginbotham L, et al. A phase II study repurposing atomoxetine for neuroprotection in mild cognitive impairment. Brain. 2022;145:1924–1938.

40. Emre M, Aarsland D, Brown R, Burn DJ, Duyckaerts C, Mizuno Y, et al. Clinical diagnostic criteria for dementia associated with Parkinson’s disease. Movement Disorders. 2007;22:1689–1707.

41. Sauer J-M, Ring BJ, Witcher JW. Clinical Pharmacokinetics of Atomoxetine: Clinical Pharmacokinetics. 2005;44:571–590.

42. Teichert J, Rowe JB, Ersche KD, Skandali N, Sacher J, Aigner A, et al. Determination of Atomoxetine or Escitalopram in human plasma by HPLC. Applications in Neuroscience Research Studies. Int J Clin Pharmacol Ther. 2020;58:426–438.

43. R Core Team. R: A language and environment for statistical computing. version 4.1.3. 2022.

44. Geller J, Winn MB, Mahr T, Mirman D. GazeR: A Package for Processing Gaze Position and Pupil Size Data. Behav Res. 2020;52:2232–2255.

45. Mathôt S, Fabius J, Van Heusden E, Van der Stigchel S. Safe and sensible preprocessing and baseline correction of pupil-size data. Behav Res. 2018;50:94–106.

46. Bowling AC, Hindman EA, Donnelly JF. Prosaccade errors in the antisaccade task: differences between corrected and uncorrected errors and links to neuropsychological tests. Exp Brain Res. 2012;216:169–179.

47. Cotti J, Panouilleres M, Munoz DP, Vercher J-L, Pélisson D, Guillaume A. Adaptation of reactive and voluntary saccades: different patterns of adaptation revealed in the antisaccade task. The Journal of Physiology. 2009;587:127–138.

48. Reteig LC, Knapen T, Roelofs FJFW, Ridderinkhof KR, Slagter HA. No Evidence That Frontal Eye Field tDCS Affects Latency or Accuracy of Prosaccades. Front Neurosci. 2018;12.

49. Roig AB, Morales M, Espinosa J, Perez J, Mas D, Illueca C. Pupil detection and tracking for analysis of fixational eye micromovements. Optik. 2012;123:11–15.

50. Coe B, Tomihara K, Matsuzawa M, Hikosaka O. Visual and Anticipatory Bias in Three Cortical Eye Fields of the Monkey during an Adaptive Decision-Making Task. J Neurosci. 2002;22:5081–5090.

51. Jantz JJ, Watanabe M, Everling S, Munoz DP. Threshold mechanism for saccade initiation in frontal eye field and superior colliculus. Journal of Neurophysiology. 2013;109:2767–2780.

52. Rizzo J-R, Hudson TE, Abdou A, Lui YW, Rucker JC, Raghavan P, et al. Disrupted Saccade Control in Chronic Cerebral Injury: Upper Motor Neuron-Like Disinhibition in the Ocular Motor System. Front Neurol. 2017;8.

53. Rivaud-Pechoux S, Vidailhet M, Brandel JP, Gaymard B. Mixing pro- and antisaccades in patients with parkinsonian syndromes. Brain. 2006;130:256–264.

54. Bahill AT, Clark MR, Stark L. The main sequence, a tool for studying human eye movements. Mathematical Biosciences. 1975;24:191–204.

55. Blundell J, Frisson S, Chakrapani A, Gissen P, Hendriksz C, Vijay S, et al. Oculomotor abnormalities in children with Niemann-Pick type C. Molecular Genetics and Metabolism. 2018;123:159–168.

56. Grogan JP, Sandhu TR, Hu MT, Manohar SG. Dopamine promotes instrumental motivation, but reduces reward-related vigour. eLife. 2020;9:e58321.

57. Manohar SG, Finzi RD, Drew D, Husain M. Distinct Motivational Effects of Contingent and Noncontingent Rewards. Psychol Sci. 2017;28:1016–1026.

58. Muhammed K, Dalmaijer E, Manohar S, Husain M. Voluntary modulation of saccadic peak velocity associated with individual differences in motivation. Cortex. 2020;122:198–212.

59. Lebedev S, Gelder PV, Tsui WH. Square-root relations between main saccadic parameters. 1996.

60. Gibaldi A, Sabatini SP. The saccade main sequence revised: A fast and repeatable tool for oculomotor analysis. Behav Res. 2021;53:167–187.

61. Calancie OG, Brien DC, Huang J, Coe BC, Booij L, Khalid-Khan S, et al. Maturation of Temporal Saccade Prediction from Childhood to Adulthood: Predictive Saccades, Reduced Pupil Size, and Blink Synchronization. J Neurosci. 2022;42:69–80.

62. Koohi N, Bancroft MJ, Patel J, Castro P, Akram H, Warner TT, et al. Saccadic Bradykinesia in Parkinson’s Disease: Preliminary Observations. Movement Disorders. 2021;36:1729–1731.

63. Singmann H, Bolker B, Westfall J, Aust F, Ben-Shachar MS, Højsgaard S, et al. Afex: Analysis of factorial experiments. 2020.

64. Lenth R, Singmann H, Love J, Buerkener P, Herve M. Emmeans: Estimated Marginal Means, Aka Least-Squares Means. 2019.

65. Morey RD, Rouder JN, Jamil T, Urbanek S, Forner K, Ly A. BayesFactor: Computation of Bayes factors for common designs. 2015.

66. Makowski D, Ben-Shachar MS, Lüdecke D. bayestestR: Describing Effects and their Uncertainty, Existence and Significance within the Bayesian Framework. Journal of Open Source Software. 2019;4:1541.

67. Kass RE, Raftery AE. Bayes Factors. Journal of the American Statistical Association. 1995;90:773–795.

68. Folstein MF, Folstein SE, McHugh PR. ‘Mini-mental state’. A practical method for grading the cognitive state of patients for the clinician. J Psychiatr Res. 1975;12:189– 198.

69. Nasreddine ZS, Phillips NA, Bédirian V, Charbonneau S, Whitehead V, Collin I, et al. The Montreal Cognitive Assessment, MoCA: A Brief Screening Tool For Mild Cognitive Impairment. Journal of the American Geriatrics Society. 2005;53:695–699.

70. Mioshi E, Dawson K, Mitchell J, Arnold R, Hodges JR. The Addenbrooke’s Cognitive Examination Revised (ACE-R): a brief cognitive test battery for dementia screening. International Journal of Geriatric Psychiatry. 2006;21:1078–1085.

71. Goetz CG, Tilley BC, Shaftman SR, Stebbins GT, Fahn S, Martinez-Martin P, et al. Movement Disorder Society-sponsored revision of the Unified Parkinson’s Disease Rating Scale (MDS-UPDRS): Scale presentation and clinimetric testing results. Movement Disorders. 2008;23:2129–2170.

72. Hutton SB, Ettinger U. The antisaccade task as a research tool in psychopathology: A critical review. Psychophysiology. 2006;43:302–313.

73. Schaeffer DJ, Chi L, Krafft CE, Li Q, Schwarz NF, McDowell JE. Individual differences in working memory moderate the relationship between prosaccade latency and antisaccade error rate. Psychophysiology. 2015;52:605–608.

74. Rayner K. Eye movements in reading and information processing: 20 years of research. Psychological Bulletin. 1998;124:372–422.

75. Robinson DA. The mechanics of human saccadic eye movement. J Physiol. 1964;174:245–264.

76. Carpenter RHS. Oculomotor Procrastination. In D. F. Fisher, R. A. Monty, & J. W. Senders (Eds.), Eye movements: Cognition and visual perception, Hillsdale: Lawrence Erlbaum; 1981. p. (pp. 237-246).

77. Findlay JM, Walker R. A model of saccade generation based on parallel processing and competitive inhibition. Behavioral and Brain Sciences. 1999;22:661–674.

78. Chamberlain SR, Müller U, Blackwell AD, Clark L, Robbins TW, Sahakian BJ. Neurochemical Modulation of Response Inhibition and Probabilistic Learning in Humans. Science. 2006;311:861–863.

79. Robinson ESJ, Eagle DM, Mar AC, Bari A, Banerjee G, Jiang X, et al. Similar Effects of the Selective Noradrenaline Reuptake Inhibitor Atomoxetine on Three Distinct Forms of Impulsivity in the Rat. Neuropsychopharmacol. 2008;33:1028–1037.

80. McBurney-Lin J, Vargova G, Garad M, Zagha E, Yang H. The locus coeruleus mediates behavioral flexibility. Cell Reports. 2022;41.

81. McGaughy J, Ross RS, Eichenbaum H. Noradrenergic, but not cholinergic, deafferentation of prefrontal cortex impairs attentional set-shifting. Neuroscience. 2008;153:63–71.

82. Tait DS, Brown VJ, Farovik A, Theobald DE, Dalley JW, Robbins TW. Lesions of the dorsal noradrenergic bundle impair attentional set-shifting in the rat. European Journal of Neuroscience. 2007;25:3719–3724.

83. Bari A, Aston-Jones G. Atomoxetine modulates spontaneous and sensory-evoked discharge of locus coeruleus noradrenergic neurons. Neuropharmacology. 2013;64:53– 64.

84. Di Stasi LL, Catena A, Cañas JJ, Macknik SL, Martinez-Conde S. Saccadic velocity as an arousal index in naturalistic tasks. Neuroscience & Biobehavioral Reviews. 2013;37:968–975.

85. Wang CA, White B, Munoz DP. Pupil-linked Arousal Signals in the Midbrain Superior Colliculus. Journal of Cognitive Neuroscience. 2022;34:1340–1354.

86. Joshi S, Li Y, Kalwani RM, Gold JI. Relationships between Pupil Diameter and Neuronal Activity in the Locus Coeruleus, Colliculi, and Cingulate Cortex. Neuron. 2016;89:221–234.

87. Coe BC, Munoz DP. Mechanisms of saccade suppression revealed in the anti-saccade task. Philosophical Transactions of the Royal Society B: Biological Sciences. 2017;372:20160192.

88. Everling S, Johnston K. Control of the superior colliculus by the lateral prefrontal cortex. Philosophical Transactions of the Royal Society B: Biological Sciences. 2013;368:20130068.

89. Johnston K, Everling S. Monkey Dorsolateral Prefrontal Cortex Sends Task-Selective Signals Directly to the Superior Colliculus. J Neurosci. 2006;26:12471–12478.

90. Cools R, Arnsten AFT. Neuromodulation of prefrontal cortex cognitive function in primates: the powerful roles of monoamines and acetylcholine. Neuropsychopharmacology. 2022;47:309–328.

91. Gamo NJ, Wang M, Arnsten AFT. Methylphenidate and Atomoxetine Enhance Prefrontal Function Through α2-Adrenergic and Dopamine D1 Receptors. Journal of the American Academy of Child & Adolescent Psychiatry. 2010;49:1011–1023.

92. Rowe JB, Hughes L, Ghosh BCP, Eckstein D, Williams-Gray CH, Fallon S, et al. Parkinson’s disease and dopaminergic therapy--differential effects on movement, reward and cognition. Brain. 2008;131:2094–2105.

93. Rae CL, Nombela C, Rodrıguez PV, Ye Z, Hughes LE, Jones PS, et al. Atomoxetine restores the response inhibition network in Parkinson’s disease. 2016;139:2235–2248.

94. Briand KA, Strallow D, Hening W, Poizner H, Sereno AB. Control of voluntary and reflexive saccades in Parkinson’s disease. Exp Brain Res. 1999;129:38–48.

95. Harris CM, Wolpert DM. The Main Sequence of Saccades Optimizes Speed-accuracy Trade-off. Biol Cybern. 2006;95:21–29.

96. Beers RJ van. Saccadic Eye Movements Minimize the Consequences of Motor Noise. PLOS ONE. 2008;3:e2070.

97. Goossens HHLM, Opstal AJ van. Optimal Control of Saccades by Spatial-Temporal Activity Patterns in the Monkey Superior Colliculus. PLOS Computational Biology. 2012;8:e1002508.

98. Goossens HHLM, Van Opstal AJ. Dynamic Ensemble Coding of Saccades in the Monkey Superior Colliculus. Journal of Neurophysiology. 2006;95:2326–2341.

99. Van Opstal JA. Neural encoding of instantaneous kinematics of eye-head gaze shifts in monkey superior Colliculus. Commun Biol. 2023;6:1–17.

100. Sparks DL. The brainstem control of saccadic eye movements. Nat Rev Neurosci. 2002;3:952–964.

101. Kardamakis AA, Moschovakis AK. Optimal Control of Gaze Shifts. J Neurosci. 2009;29:7723–7730.

102. Connolly AJ, Rinehart NJ, Johnson B, Papadopoulos N, Fielding J. Voluntary saccades in attention-deficit/hyperactivity disorder: Looking into the relationship between motor impairment and Autism Spectrum Disorder symptoms. Neuroscience. 2016;334:47–54.

103. Federighi P, Ramat S, Rosini F, Pretegiani E, Federico A, Rufa A. Characteristic Eye Movements in Ataxia-Telangiectasia-Like Disorder: An Explanatory Hypothesis. Frontiers in Neurology. 2017;8.

104. Federighi P, Cevenini G, Dotti MT, Rosini F, Pretegiani E, Federico A, et al. Differences in saccade dynamics between spinocerebellar ataxia 2 and late-onset cerebellar ataxias. Brain. 2011;134:879–891.

105. Rosini F, Federighi P, Pretegiani E, Piu P, Leigh RJ, Serra A, et al. Ocular-Motor Profile and Effects of Memantine in a Familial Form of Adult Cerebellar Ataxia with Slow Saccades and Square Wave Saccadic Intrusions. PLOS ONE. 2013;8:e69522.

106. Zaino D, Serchi V, Giannini F, Pucci B, Veneri G, Pretegiani E, et al. Different saccadic profile in bulbar versus spinal-onset amyotrophic lateral sclerosis. Brain. 2023;146:266–277.

107. Bymaster FP, Katner JS, Nelson DL, Hemrick-Luecke SK, Threlkeld PG, Heiligenstein JH, et al. Atomoxetine Increases Extracellular Levels of Norepinephrine and Dopamine in Prefrontal Cortex of Rat: A Potential Mechanism for Efficacy in Attention Deficit/Hyperactivity Disorder. Neuropsychopharmacology. 2002;27:699– 711.

108. Swanson CJ, Perry KW, Koch-Krueger S, Katner J, Svensson KA, Bymaster FP. Effect of the attention deficit/hyperactivity disorder drug atomoxetine on extracellular concentrations of norepinephrine and dopamine in several brain regions of the rat. Neuropharmacology. 2006;50:755–760.

109. Lu Z, Buchanan T, Kennard C, FitzGerald JJ, Antoniades CA. The effect of levodopa on saccades – Oxford Quantification in Parkinsonism study. Parkinsonism & Related Disorders. 2019;68:49–56.

110. Waldthaler J, Stock L, Student J, Sommerkorn J, Dowiasch S, Timmermann L. Antisaccades in Parkinson’s Disease: A Meta-Analysis. Neuropsychol Rev. 2021;31:628–642.

