## Supplementary Material for "Noradrenergic modulation of saccades in Parkinson’s disease"


1. Mood and behaviour questionnaires Page 2

2. Within session physiological effects Page 4

3. Within session subjective effects Page 6

4. Recording and equipment details Page 8

5. Prosaccade velocity and amplitude over time Page 9

6. Relating disease severity and cognitive impairment with saccades Page 10

7. References Page 11

### 1. Mood and behaviour questionnaires

Individuals with Parkinson's disease completed self-rated questionnaires to assess mood and behavioural symptoms. These included assessment of anxiety and depression (Hospital Anxiety and Depression scale; HADS) [1], impulsivity (Barratt Impulsiveness Scale; BIS-11[2], apathy (Apathy Scale [3]; Motivation and Energy Inventory; MEI) [4] and REM sleep behaviour disorder (REM sleep behaviour disorder screening questionnaire; RBDSQ [5]. Controls also completed all of these self-rated questionnaires, apart from the RBDSQ. Informant-rated questionnaires were collected from a relative or friend of the patients. These included informant versions of the AS and a general mood and behaviour symptom inventory (Cambridge Behavioural Inventory Revised; CBI-R [6].

The groups for the most part did not differ on self-reported questionnaires, with the only significant difference being lower scores in the patient group for the physical subscale of the Motivation and Energy Inventory (Supplementary Table 1).

*Supplementary Table 1*

| **Measure** | | **PD** | **Controls** | **$BF_{10}$** | ***p*** |
|---|---|---|---|---|---|
| Apathy Scale | Total Score (self-rated) | 12.68 (5.77) | 10.6 (5.2) | 0.56 | .224 |
| | Total Score (informant-rated) | 13.13 (5.59) | | | |
| BIS | Total Score | 56.45 (10.34) | 56.06 (9.86) | 0.3 | .907 |
| | Attention | 14.16 (4.3) | 14.16 (3.78) | 0.3 | .999 |
| | Motor | 20.08 (2.65) | 20.96 (3.4) | 0.42 | .353 |
| | Non-planning | 22.21 (5.4) | 20.94 (4.49) | 0.39 | .430 |
| HADS | Anxiety | 4.53 (3.2) | 4.36 (3.4) | 0.3 | .882 |
| | Depression | 3.95 (2.68) | 2.8 (2.78) | 0.64 | .174 |
| MEI | Total Score | 98.05 (21.3) | 108.48 (16.87) | 1.10 | .088 |
| | Mental | 44.11 (8.97) | 47.28 (8.25) | 0.54 | .236 |
| | Physical | 23.95 (6.95) | 29.16 (5.96) | **4.71** | **.013** |
| | Social | 30 (7.34) | 32.04 (5.29) | 0.48 | .313 |
| RBDSQ | | 4.58 (3.45) | | | |
| CBI | Total Score | 15.13 (13.6) | | | |
| | Abnormal Behaviour | 0.84 (1.12) | | | |
| | Beliefs | 0.37 (1.21) | | | |
| | Eating Habits | 0.95 (1.58) | | | |
| | Everyday Skills | 1.16 (2.41) | | | |
| | Memory and Orientation | 4.66 (3.9) | | | |
| | Mood | 1.26 (2.1) | | | |
| | Motivation | 2.26 (3.35) | | | |
| | Stereotypic and Motor Behaviours | 0.79 (1.4) | | | |
| | Self Care | 0.42 (0.84) | | | |
| | Sleep | 2.42 (2.17) | | | |

*Note*: Data are presented as mean (SD). Group comparisons were performed with independent samples t-tests. $BF_{10}$, default Bayes Factor for the alternative hypothesis versus the null hypothesis; *p*, two-tailed *p*-values, uncorrected for multiple comparisons.

### 2. Within session physiological effects

As described below, there was evidence of increased pulse rates under atomoxetine, when assessed in the upright (but not the supine) position. These were not considered clinically significant. Systolic and diastolic blood pressure was also increased under atomoxetine for supine (but not upright) measures. There was evidence for a time effect on supine blood pressure measures, where they were raised at completion of testing, relative to the measures on arrival and two-hours post tablet administration. Mean values and ranges for blood pressure and pulse rates are shown in Supplementary Table 2.

*Pulse rates*

Supine/Lying down pulse rates did not change significantly under atomoxetine vs. placebo, as evidenced by a lack of main effect ($F_{(1, 89)} = 5.57$, $p = .020$; BF = 1.91), and did not vary across the three time points (i.e., arrival, two-hours post tablet, on completion of testing; $F_{(2, 89)} = 3.00$, $p = .055$; BF = 0.90). Upright pulse rates were increased under atomoxetine, showing a significant main effect ($F_{(1, 88.03)} = 20.99$, $p < .001$; BF = 629.81), and a significant interaction between drug status and time point ($F_{(2, 88.03)} = 6.43$, $p = .002$; BF = 13.75) driven by higher pulse rates under atomoxetine at two hours post administration ($t_{(88)} = 10.93$, $p < .001$, BF = 4.56) and on completion of testing ($t_{(88)} = 10.39$, $p < .001$, BF = 6.57).

*Blood pressure*

Supine systolic and diastolic blood pressure was raised under atomoxetine, as evidenced by significant main effects (systolic: $F_{(1, 89)} = 9.86$, $p = .002$; BF = 13.39; diastolic: $F_{(1, 89)} = 16.21$, $p < .001$; BF = 163.03). Upright systolic and diastolic blood pressure did not show an overall change under atomoxetine, as evidenced by the lack of main effects (systolic: $F_{(1, 88)} = 2.41$, $p = .124$; BF = 0.57; diastolic: $F_{(1, 88)} = 0.00$, $p = .972$; BF = 0.19). However, there was a main effect of time point (systolic: $F_{(1, 88)} = 5.86$, $p = .004$; BF = 8.47; diastolic: $F_{(1, 88)} = 5.98$, $p = .004$; BF = 8.24), driven by increased blood pressure on completion of testing, compared to arrival and two hours post (systolic, arrival vs. completion: $t_{(88)} = 9.77$, $p = .011$, BF = 1.94; systolic, two hours post vs. completion: $t_{(88)} = 9.79$, $p = .010$, BF = 3.52; diastolic, arrival vs. completion: $t_{(88)} = 4.87$, $p = .012$, BF = 2.30; diastolic, two hours post vs. completion: $t_{(88)} = 5.60$, $p = .006$, BF = 3.99).

*Supplementary Table 2*

| **Measure** | | | **Placebo** | **Atomoxetine** |
|---|---|---|---|---|
| Pulse rates | Lying | Arrival | 70.00 (33.5 – 55.5; 8.85) | 70.58 (56 – 91; 10.27) |
| | | 2-hours | 69.95 (55 – 86; 10.37) | 74.89 (58 – 95; 11.25) |
| | | Completion | 66.95 (50 – 85; 8.20) | 70.21 (50 – 93; 11.06) |
| | Upright | Arrival | 75.63 (49 – 100; 12.64) | 74.95 (56 – 110; 14.30) |
| | | 2-hours | 70.00 (54 – 86; 8.88) | 80.95 (60 – 106; 13.91) |
| | | Completion | 68.33 (49 – 80; 7.88) | 79.95 (57 – 116; 15.39) |
| Systolic blood pressure | Lying | Arrival | 127.68 (84 – 151; 16.67) | 133.26 (109 – 175; 17.24) |
| | | 2-hours | 125.32 (95 – 156; 15.03) | 135.11 (114 – 169; 15.35) |
| | | Completion | 131.21 (116 – 155; 12.54) | 136.58 (83 – 186; 24.03) |
| | Upright | Arrival | 124.37 (80 – 165; 21.78) | 130.26 (90 – 166; 21.20) |
| | | 2-hours | 123.21 (101 – 145; 12.47) | 131.37 (93 – 167; 18.68) |
| | | Completion | 139.11 (118 – 176 14.60) | 136.11 (97 – 183; 20.99) |
| Diastolic blood pressure | Lying | Arrival | 70.89 (50 – 84; 8.61) | 74.00 (55 – 85; 7.46) |
| | | 2-hours | 68.37 (54 – 80; 6.68) | 74.53 (58 – 94; 9.04) |
| | | Completion | 72.42 (59 – 87; 7.07) | 77.21 (63 – 98; 9.93) |
| | Upright | Arrival | 72.89 (43 – 82; 9.71) | 73.32 (56 – 86; 8.09) |
| | | 2-hours | 71.26 (51 – 91; 10.78) | 73.47 (52 – 93; 9.82) |
| | | Completion | 79.44 (61 – 93; 8.15) | 76.68 (61 – 92; 9.08) |

*Note*: Data are presented as mean (range; SD).

#### 3. Within session subjective effects

Although the visual analogue scale (VAS) is a continuous measure, participants often respond at either end of the scale, leading to bi- or even tri-modal distributions (Supplementary Figure 6). Such dynamics are not well captured by conventional analyses (e.g., linear regression) that assume multivariate normality. To address this issue, we analysed the VAS data using a Bayesian ordered beta regression model [7]. The strength of this model is that it simultaneously estimates the probability of responses at the scale's lower and upper bounds as well as continuously distributed responses in between the bounds. As described below, there was no change in subjective mood/arousal levels within the sessions.

*Supplementary Figure 1*

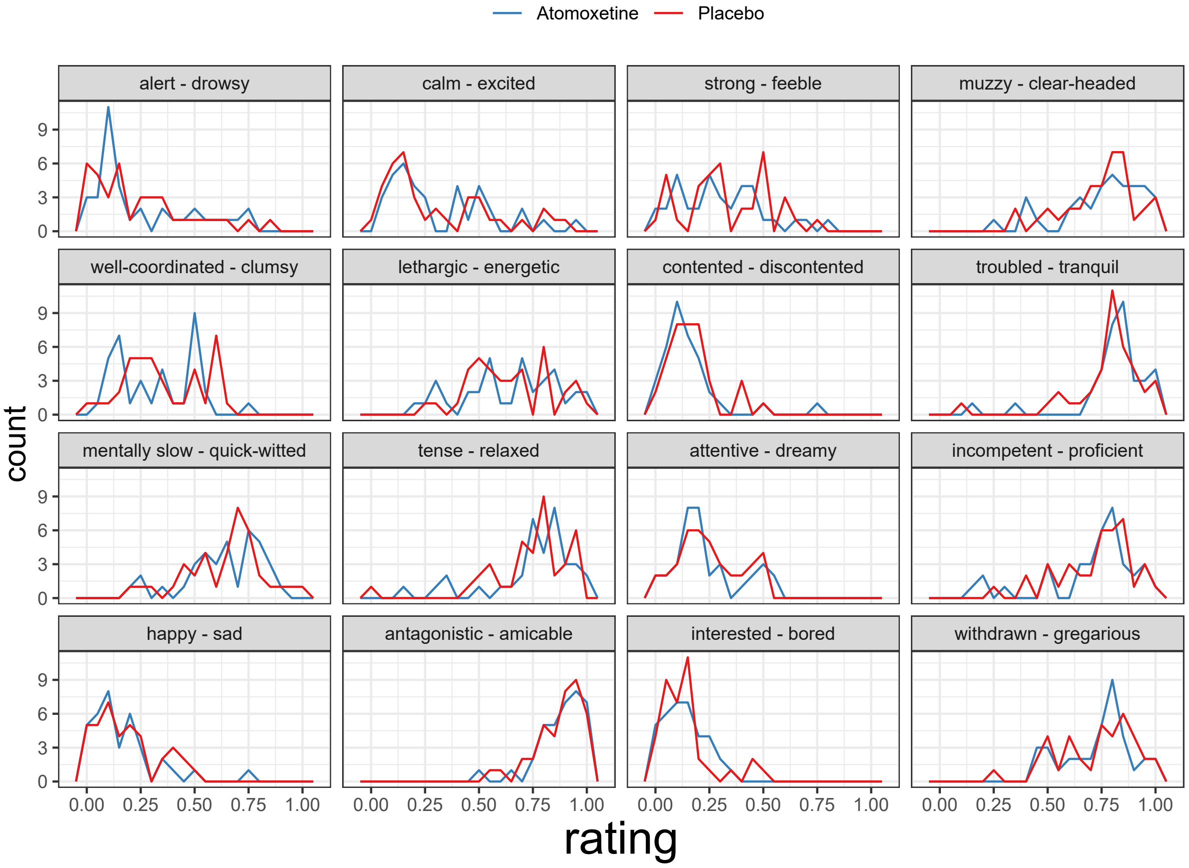


Frequency polygons of VAS ratings in the PD group. Each panel represents one VAS item, as described by the panel titles. The first term of each panel title corresponds to the left extreme of that VAS item (i.e. rating = 0), whereas the second term corresponds to the right extreme (i.e. rating = 1).

We modelled drug (atomoxetine vs. placebo), time point (2 hours post administration vs. baseline), the drug × time interaction, and session (first vs. second visit) as categorical predictors of the VAS response (i.e., fixed effects), and we allowed the intercept to vary by VAS item and by participant (i.e., random effects). Following Kubinec (2020), we assigned a weakly informative normal prior on the regression coefficients: $\beta \sim N(0, 5)$. For posterior inference, we set a region of practical equivalence (ROPE) at $\pm 0.1 \times SD_{VAS} = \pm 0.019$, corresponding to a negligible effect size [8,9].

There were no main effects of drug or time point on VAS response, as the posterior distributions of these coefficients were largely contained by the ROPE (Supplementary Figure 7; drug: β = -0.02, 95% HDI [-0.06, 0.03], proportion in ROPE = 46.20%; time point: β = 0.02, 95% HDI [-0.03, 0.06], proportion in ROPE = 49.69%). Although the posterior estimate of the drug × time point interaction effect was greater than the upper bound of the ROPE, we failed to reject the null as a relatively large proportion of the posterior distribution was contained by the ROPE (interaction: β = 0.03, 95% HDI [-0.01, 0.08], proportion in ROPE = 24.01%). Taken together, these results suggest that atomoxetine did not induce a significant change in subjective states, as measured by the VAS.

*Supplementary Figure 2*

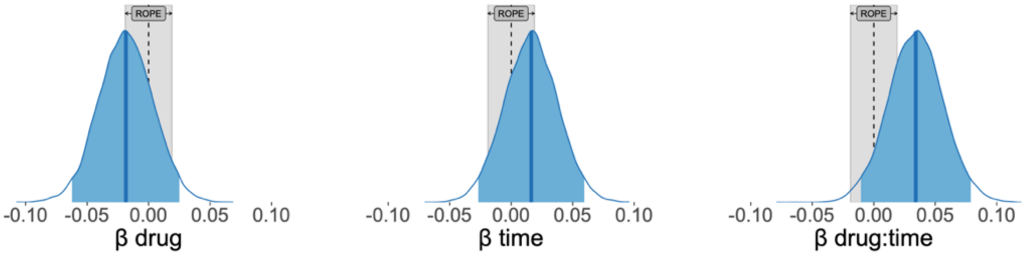


Posterior distributions of predictors of VAS responses. For each panel, the dark blue vertical line represents the median – that is, the posterior estimate of the regression coefficient; the blue shaded area represents the 95% highest density interval of the posterior distribution; and the blue density trace represents the full posterior distribution. The grey area represents a region of practical equivalence (ROPE), corresponding to a negligible effect size of ±0.1.

### 4. Recording and equipment details

Pupil size and eye movements were recorded using an EyeLink-1000 portable duo eye tracker (SR Research, Osgoode, ON, Canada) at a sampling rate of 500 Hz. Head movement was restricted using a fixed chin rest and 3-point horizontal calibration was conducted prior to each task. Testing took place in a well-lit room, with light levels kept constant across sessions. Stimuli were displayed on a 15-inch laptop screen (1920 × 1080-pixel) placed 55 cm directly in front of the participant. Our analyses used saccade detection, velocity and amplitude calculations from the standard Eyelink algorithms. The tasks were programmed using Eyelink Experiment Builder software, with further processing and statistical analyses conducted in R (version 4.2.1, R Core Team, 2022).

### 5. Prosaccade velocity and amplitude over time

In the Parkinson's disease group, prosaccade mean velocity residuals across 10 time bins were not significantly different after atomoxetine vs. placebo (Figure 1a; $F(1, 16.74) = 0.03$, $p = 0.873$, BF = 0.145). As expected, given the typical saccade trajectory, there was a main effect of time bin ($F(9, 163.18) = 127.43$, $p = < 0.001$, BF = $1.24 \times 10^{64}$), with no interaction between drug condition and time bin ($F(9, 152.71) = 1.01$, $p = 0.434$, BF = 0.050). There was no significant effect of visit order ($F(1, 16.71) = 1.53$, $p = 0.233$, BF = 0.242).

Amplitude across the 10 time points did not differ significantly between the atomoxetine and placebo conditions (Figure 1b; $F(1, 16.75) = 1.13$, $p = 0.304$, BF = 0.181). There was a significant main effect of time bin ($F(9, 164.01) = 439.45$, $p < 0.001$, BF = $1.59 \times 10^{104}$), with no interaction between drug condition and time bin ($F(9, 143.31) = 0.69$, $p = 0.714$, BF = $4.84 \times 10^{-6}$) and no effect of visit ($F(1, 16.94) = 0.23$, $p = 0.635$, BF = 0.133).

Comparing the Parkinson's disease group on placebo with control participants, velocity was not significantly different ($F(1, 41.80) = 0.001$, $p = 0.973$, BF = 0.147). There was a main effect of time bin ($F(9, 372.81) = 140.50$, $p < 0.001$, BF = $5.98 \times 10^{111}$), with no interaction between group and time bin ($F(9, 372.81) = 0.96$, $p = 0.476$, BF = 0.003). Amplitude was not significantly different between groups ($F(1, 41.66) = 1.30$, $p = 0.261$, BF = 0.387); there was a significant effect of time bin ($F(9, 367.68) = 519.32$, $p < 0.001$, BF = $5.81 \times 10^{204}$) with no interaction between group and time bin ($F(9, 367.68) = 0.58$, $p = 0.814$, BF = $3.46 \times 10^{-4}$).

### 6. Relating disease severity and cognitive impairment with saccades

In the main text, we tested whether prosaccade latency, antisaccade error rate or main sequence deviation (i.e., Δ peak velocity values from controls' main sequence) in the placebo group correlated with global cognition (ACE-R total score) or disease severity (MDS-UPDRS-III score). We conducted Pearson correlations with FDR correction applied within both families of comparisons. For global cognition we found no significant relationships (latency: r = -0.256, $p_{(adjust)}$ = 0.290, BF = 0.752; error rate: r = -0.282, $p_{(adjust)}$ = 0.290, BF = 0.813; Δ peak velocity: r = -0.323, $p_{(adjust)}$ = 0.290, BF = 0.986). For disease severity, we found no significant relationship for antisaccade error rate (r = 0.023, $p_{(adjust)}$ = 0.928, BF = 0.497), but both prosaccade latency and main sequence deviation showed a positive correlation consistent with worsening impairment with disease progression (latency: r = 0.549, $p_{(adjust)}$ = 0.022, BF = 4.81; Δ peak velocity: r = 0.692, $p_{(adjust)}$ = 0.003, BF = 29.89). See Supplementary Figure 3 below.

*Supplementary Figure 3*

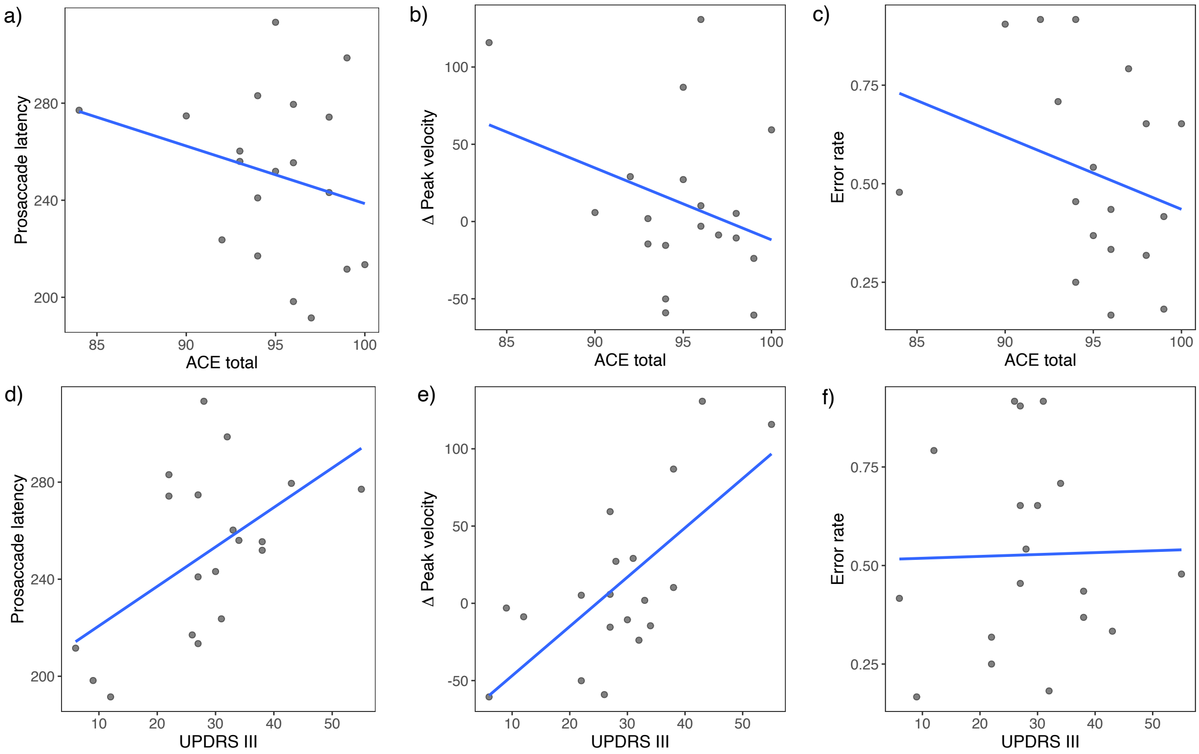


For Parkinson's disease on placebo, plots showing relationship between ACE total score with a) prosaccade latency, b) Δ peak velocity and c) antisaccade error rate; MDS-UPDRS-III score with d) prosaccade latency, e) Δ peak velocity and f) antisaccade error rate.

### 7. References


1. Zigmond AS, Snaith RP. The Hospital Anxiety and Depression Scale. Acta Psychiatrica Scandinavica. 1983;67:361–370.
2. Patton JH, Stanford MS, Barratt ES. Factor structure of the barratt impulsiveness scale. Journal of Clinical Psychology. 1995;51:768–774.
3. Starkstein SE, D P, Mayberg HS, D TPM, A PAM, D RLM, et al. Reliability, validity, and clinical correlates of apathy in Parkinson's disease. Journal of Neuropsychiatry and Clinical Neurosciences. 1992:134.
4. Fehnel SE, Bann CM, Hogue SL, Kwong WJ, Mahajan SS. The development and psychometric evaluation of the motivation and energy inventory (MEI). Qual Life Res. 2004;13:1321–1336.
5. Stiasny-Kolster K, Mayer G, Schäfer S, Möller JC, Heinzel-Gutenbrunner M, Oertel WH. The REM sleep behavior disorder screening questionnaire—A new diagnostic instrument. Movement Disorders. 2007;22:2386–2393.
6. Wear HJ, Wedderburn CJ, Mioshi E, Williams-Gray CH, Mason SL, Barker RA, et al. The Cambridge Behavioural Inventory revised. Dementia & Neuropsychologia. 2008;2:102–107.
7. Kubinec R. Ordered Beta Regression: A Parsimonious, Well-Fitting Model for Survey Sliders and Visual Analog Scales. SocArXiv. 2020. 2 March 2020. https://doi.org/10.31235/osf.io/2sx6y.
8. Cohen J. Statistical Power Analysis for the Behavioral Sciences. New York: Routledge; 1988.
9. Kruschke JK. Rejecting or Accepting Parameter Values in Bayesian Estimation. Advances in Methods and Practices in Psychological Science. 2018;1:270–280.
10. R Core Team. R: A language and environment for statistical computing. version 4.1.3. 2022.
